# Effectiveness of digital contact tracing for COVID-19 in New South Wales, Australia

**DOI:** 10.1101/2021.11.18.21266558

**Authors:** Florian Vogt, Bridget Haire, Linda Selvey, John Kaldor

## Abstract

**Background:** Digital proximity tracing applications were rolled out early in the COVID-19 pandemic in many countries to complement conventional contact tracing. Empirical evidence about their benefits for pandemic response remains scarce. We evaluated the effectiveness and usefulness of ‘*COVIDSafe*’, Australia’s national smartphone-based proximity tracing application for COVID-19.

**Methods:** In this prospective study, conducted in New South Wales, Australia between May and November 2020, we calculated the positive predictive value and sensitivity of *COVIDSafe*, its additional contact yield, and the number of averted public exposure events. Semi-structured interviews with public health staff were conducted to assess the application’s usefulness.

**Results:** There were 619 confirmed COVID-19 cases and over 25,300 close contacts during the study period. *COVIDSafe* was used by 137 (22%) cases and detected 79 (0·3%) close contacts. It had a positive predictive value of 39% and a sensitivity of 15%, and detected 17 (0·07%) additional close contacts that were not identified by conventional contact tracing. The application generated a substantial additional workload for public health staff and was not considered useful.

**Conclusions:** *COVIDSafe* was not sufficiently effective to make a meaningful contribution to the COVID-19 response in Australia’s most populous state over a 6-month period. This contrasts optimistic projections from modelling studies about the added value of digitally supported contact tracing. We found no evidence that it adds value to conventional contact tracing, and recommend that their implementation should always include comprehensive effectiveness evaluations.

## BACKGROUND

Contact tracing is a core component of the public health response to the Coronavirus 2019 (COVID-19) pandemic.^(1)^ It aims to interrupt transmission chains by identifying people who had contact with a case so that those at risk of infection can be promptly quarantined, thereby reducing further transmission. Every newly diagnosed case is interviewed by public health staff to identify contacts during their infectious period. Contacts who are assessed to be at high risk of infection are then notified and directed to quarantine. ^(2)^ This conventional approach to contact identification and notification is time-consuming and resource-intensive, and can become quickly overwhelmed when incidence is high.

Smartphone-based proximity tracing applications were viewed with considerable enthusiasm early in the COVID-19 pandemic as a means of overcoming limitations of conventional contact tracing,^(3)^ and a wide range of applications were quickly rolled out.^(4-6)^ Most digital contact tracing applications use Bluetooth technology, wherein the occurrence of a ‘contact’ between two smartphone users is indicated by the duration, frequency and transmission strength of Bluetooth signal exchanges.^(7)^ Information is either stored on individuals’ phones for a determined period of time (decentralised approach) or uploaded into a common database (centralised approach).^(3)^ Under the decentralised model, adopted in Canada, Finland, Germany, Switzerland, the United Kingdom and Vietnam, the matching of contact information occurs on people’s phones without involvement of public health authorities, and contact notification usually occurs automatically. By contrast, contact identification under the centralised approach occurs via a common database, which gives public health authorities access to undertake contact risk assessment and notification.^(3)^ This approach has been implemented by Australia, China, France, New Zealand, Singapore and Taiwan.^(8-12)^

Despite implementation in many countries, little empirical evidence exists about the effectiveness of digital tracing applications.^(13, 14)^ While two recent evaluations from the United Kingdom and Switzerland suggest that systems based on the decentralised approach may be able to provide some benefits to a country’s COVID-19 response under certain conditions,^(15, 16)^ no evidence exists to date about the population-level effectiveness of decentralized digital contact tracing applications. Considering the substantial investments to develop, promote and maintain these systems, it is important to establish how well they work, and what their added value within the existing contact tracing system is.^(17)^ Therefore, we assessed the effectiveness and usefulness of the *COVIDSafe* app, Australia’s national smartphone-based proximity tracing application.

## METHODS

### Design and objectives

A prospective study was conducted to assess the *COVIDSafe* app (hereinafter ‘the app’) as an integrated tool within the existing contact tracing system in the state of New South Wales (NSW), Australia between May and November 2020. Our objectives were to assess the effectiveness of the app to detect close contacts and prevent public exposure events, and its usefulness during the contact identification and risk assessment process.

### The use of the COVIDSafe app in New South Wales

In Australia, public health is the responsibility of state and territory governments, with the national government providing funding and coordination. The national government launched the *COVIDSafe* app on 26 April 2020 with the goal of enhancing COVID-19 contact tracing nationwide. The app is based on the centralised approach, and is intended to supplement rather than to replace conventional interview-based contact tracing. Each app user’s smartphone stores coded information communicated by other smartphones that have come in sufficiently close proximity to exchange Bluetooth signals. These data are automatically deleted after a rolling 21-day period. Once a person is diagnosed with SARS-CoV-2 infection, public health staff can seek informed consent from the case to access app data for contact tracing purposes. App use is ascertained as part of the standard case interview. If consent is provided, data are uploaded to the *COVIDSafe* National Data Store database. Data include names or user-selected aliases, phone numbers, postcodes of residence, age ranges, and dates and times of signal exchanges between phones. For privacy reasons, no information is collected about the location where signal exchanges took place. The database also contains information on the strength and length of the exchanged Bluetooth signals, which allows estimation of the duration and physical proximity between phones during the signal exchange. An in-built filtering algorithm selects only those contacts whose signal exchange(s) with the case’s smartphone meet pre-defined thresholds. These thresholds were set to approximate the national close contact definition in use at that time, namely face-to-face contact for at least 15 minutes over the course of one week during the case’s infectious period or presence in an enclosed space with an infectious case for at least two hours.^(18)^ This list of app-suggested contacts is accessible to public health staff via a secure web portal, who then undertake a risk assessment and reconciliation process of app data with information obtained from the case interview and from other sources such as attendance lists of venues or institutions. At the end of this risk assessment process, only those app-suggested contacts who are considered by public health staff to meet the close contact definition criteria are contacted and asked to self-quarantine for 14 days from the date of exposure.

The web portal access became available to NSW public health authorities on 8 May 2020. Between May and July 2020, the national Department of Health and the Digital Transformation Agency worked together with state and territory health authorities, including NSW, to improve the utility of the app and the web portal. During this period, app-related public health activities in NSW were managed by the Public Health Response Branch within the NSW Ministry of Health. From August 2020 onwards, the app was progressively rolled out to public health units (PHUs) in NSW, situated within each local health district of the state.

### Study population and data collection

We included all confirmed SARS-CoV-2 cases notified in NSW between 4 May and 4 November 2020 unless they were in quarantine during their entire infectious period. Cases aged 12 years and younger were excluded since app use was not systematically assessed for this age group. Persons with overseas-acquired infections were also excluded since all international arrivals into NSW were required to complete 14 days in state-managed hotel quarantine directly upon arrival. A case was defined as app-using if they reported having the app installed and running on their smartphone for at least part of their infectious period. We extracted information on app-using cases from the NSW Notifiable Conditions Information Management System (NCIMS), which is used to record standardized information for all confirmed SARS-CoV-2 cases in NSW. In addition, we maintained a dedicated database during the study period to track app-related parameters for app-using cases and their contacts. Information collected included whether app data were accessed by public health staff, the reason for not accessing app data where applicable, the total number of app-suggested contacts during the case’s infectious period, and the number of app-suggested contacts determined by the public health staff to be close contacts based on the risk assessment and data reconciliation process. App use was recorded for close contacts if they were app-suggested.

Additionally, we conducted six semi-structured interviews with public health staff in NSW to assess the app’s usefulness in the contact tracing process. Public health staff at PHU level with experience in using the app were initially invited via a group email and subsequently followed up individually. Five interviews were conducted through video conferencing platforms, audio recorded and transcribed verbatim, while the sixth interview was conducted over email. Interviews took between 20 and 60 minutes.

### Data analysis

The demographic, socio-economic, and epidemiological characteristics of cases were stratified by app usage. Comparisons between app and non-app-using cases were assessed using chi-squared tests for categorical variables, and Wilcoxon rank-sum tests for medians of continuous variables. We calculated the number and percentage of cases whose app data were accessed and used, as well as the number and percentage of close contacts generated from those cases. We also calculated the additional contact yield and the number of prevented exposure events attributable to the app. We calculated the positive predictive value of app-suggested contacts, defined as the proportion of app-suggested contacts assessed by public health staff to meet the close contact definition. We also estimated the sensitivity of the app, defined as the proportion of all close contacts identified by conventional contact tracing methods that were app-suggested. Links between surveillance data of cases and close contacts were not documented in the NCIMS before 1 August 2020, and app use among contacts that were not app-suggested was not systematically ascertained. Therefore, we estimated the number of close contacts that could be expected to have been using the app by applying the same percentage of app usage among cases to the number of close contacts that were identified by conventional contact tracing and registered in the NCIMS between 1 August and 4 November 2020. All statistical analyses were performed using STATA v.15 (StataCorp, College Station, Texas, United States). For the qualitative data analysis, we conducted thematic data analysis with data managed using NVIVO v.12 (QSR International, Melbourne, Australia).

### Ethics

We obtained ethics approval from the University of New South Wales Human Research Ethics Committee (reference number HC200468).

## RESULTS

There were 619 confirmed SARS-CoV-2 cases over the age of 12 years with infection acquired within Australia recorded in NSW between 4 May and 4 November 2020, and over 25,300 close contacts were identified through conventional contact tracing during the same period. Twenty-two percent (n=137) of cases used the app for at least part of their infectious period. App-using cases were less likely to live in socioeconomically disadvantaged areas, and more likely to be born in Australia than non-app-using cases (Table 1). They were also more likely to have acquired infection from a contact outside their household or as part of a community cluster, and to have more close contacts than cases not using the app. There were no significant differences by sex, age, or geographic remoteness.

**TABLE 1:** CHARACTERISTICS OF AUSTRALIAN-ACQUIRED SARS-COV-2 CASES IN NSW BY *COVIDSAFE* APP USE, 4 MAY TO 4 NOVEMBER 2020

|  | **App-using cases<sup>1</sup>**<br>**(N=137)** | **Cases not using the app<sup>1</sup>**<br>**(N=482)** | **p-value** |
| --- | --- | --- | --- |
| **Sex** |  |  | 0.823 |
| Female | 71 (52%) | 255 (53%) |  |
| Male | 66 (48%) | 227 (47%) |  |
| **Median age, years (range)** | 38 (13-81) | 40 (13-93) | 0.643 |
| **Remoteness of local health district of residence** |  |  | 0.102 |
| Major city local health districts | 133 (98%) | 453 (94%) |  |
| Regional local health districts | 3 (2%) | 27 (6%) |  |
| **Postcode-level index of relative social disadvantage<sup>2</sup>** |  |  | <0.001 |
| Least disadvantaged | 71 (52%) | 162 (34%) |  |
| Less disadvantaged | 40 (29%) | 128 (27%) |  |
| Most disadvantaged | 26 (19%) | 191 (39%) |  |
| **Country of birth** |  |  | 0.026 |
| Australia | 76 (55%) | 209 (43%) |  |
| Overseas | 49 (36%) | 234 (49%) |  |
| Not stated | 12 (9%) | 39 (8%) |  |
| **Likely source of infection** |  |  | 0.005 |
| Household contact | 33 (24%) | 170 (35%) |  |
| Other contact/cluster-associated | 93 (68%) | 246 (51%) |  |
| Source not determined | 9 (7%) | 45 (9%) |  |
| Interstate acquired | 2 (1%) | 20 (4%) |  |
| **Median number of close contacts<sup>3</sup> (range)** | 5 (0-121) | 2 (0-196) | <0.001 |

<sup>1</sup> Excluding missing values.
<sup>2</sup> Based on the Australian Bureau of Statistics' Index of Relative Socioeconomic Disadvantage data.
<sup>3</sup> For the time period 1 August to 4 November 2020 only, when linkage between cases and contacts was of sufficient quality to report estimates.

App data were accessed by public health staff for 92 (67%) app-using cases (Figure 1). Reasons for not accessing app data were: the case being in isolation during their entire infectious period (n=31, 69%); technical issues (n=5; 11%); loss to follow-up (n=4; 9%); and declining consent (n=3; 7%). Among those whose data were accessed, the app did not record any contact during the infectious period for 60 (65%) cases, leaving 32 with at least one app-suggested contact (5% of the total 619 cases in NSW). These 32 cases had 205 app-suggested contacts. Following the risk assessment and data reconciliation by public health staff, 79 app-suggested contacts were assessed as meeting the close contact definition and were directed to self-quarantine, resulting in a positive predictive value of the app in correctly identifying close contacts of 39% (79/205). Common examples of app-suggested contacts who did not meet the close contact definition included: neighbours in apartment buildings; office workers in adjacent rooms; customers in neighbouring restaurants; and people waiting in separate cars at COVID-19 drive-through testing clinics. An estimated 85% of close contacts identified by conventional contact tracing who could be expected to use the app were not detected by the app, resulting in an estimated sensitivity of the app to detect close contacts of 15% (35/236 between 1 August to 4 November, see Table 2).

**FIGURE 1:**
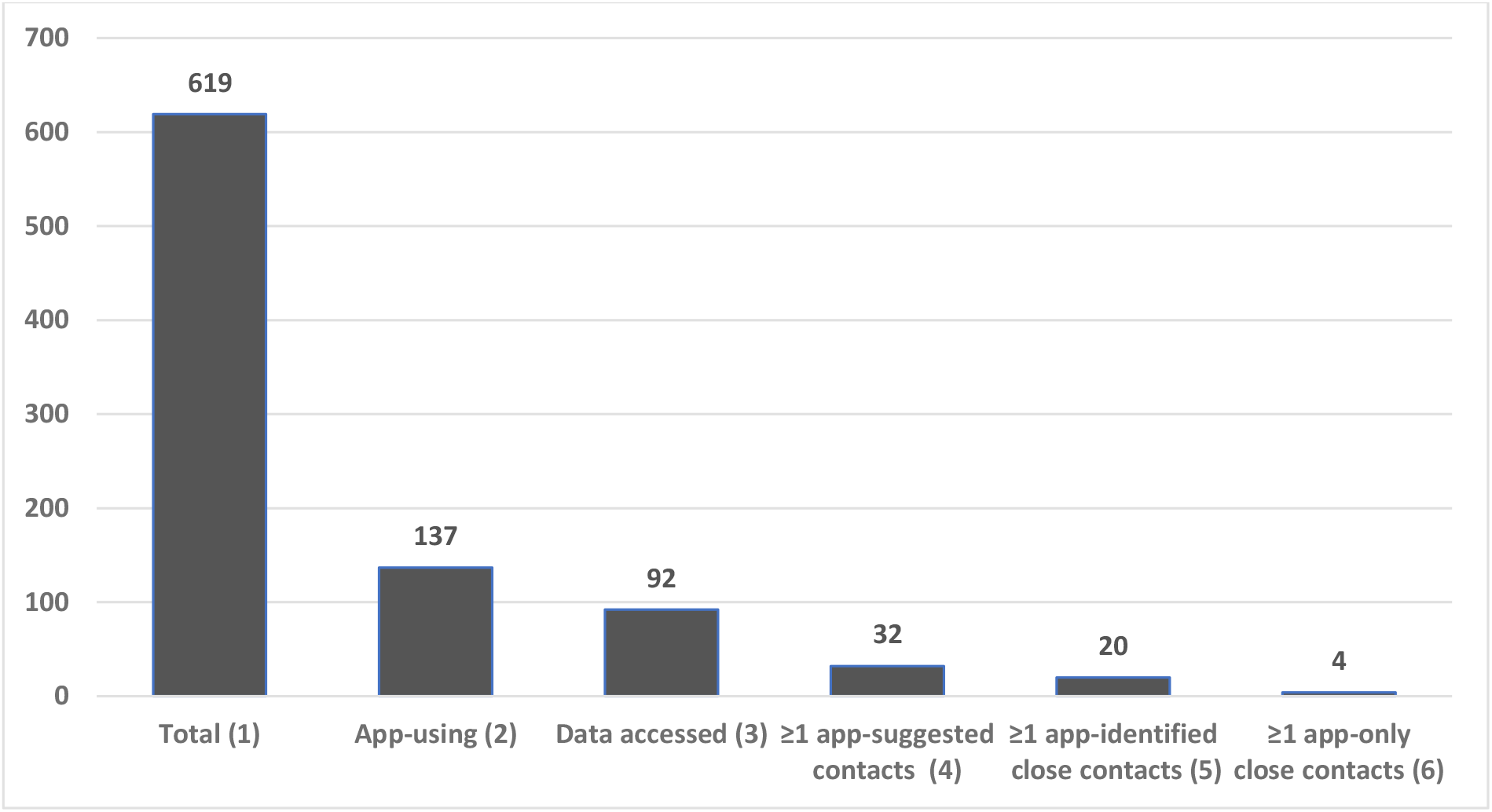
USE OF *COVIDSAFE* APP DATA FOR CONTACT TRACING AMONG SARS-COV-2 CASES IN NSW, 4 MAY TO 4 NOVEMBER 2020 (1) Total number of Australian-acquired SARS-CoV-2 cases recorded during the study period (2) Number of cases reporting using the app during infectious period (3) Number of cases whose app data were accessed by public health staff (4) Number of cases with at least one app-suggested contact (5) Number of cases with at least one app-suggested contact meeting the close contact definition (after risk assessment by public health staff) (6) Number of cases with at least one app-suggested close contact that were not also detected by conventional contact tracing.

**TABLE 2:** ESTIMATED SENSITIVITY OF THE *COVIDSAFE* APP TO DETECT CLOSE CONTACTS IN NSW, 1 AUGUST TO 4 NOVEMBER 2020

|  | Close contacts<sup>1</sup> | Not close contacts<sup>1</sup> | Total |
| --- | --- | --- | --- |
| **App-suggested contacts<sup>2</sup>** | 35 | NA | NA |
| **Not app-suggested contacts<sup>3</sup>** | 200 | NA | NA |
| **Total app-using contacts<sup>3</sup>** | 236 | NA | NA |

During the period 1 August to 4 November 2020, when data of cases and contacts could be linked, app-using cases had a total of 1,073 close contacts identified either via conventional contact tracing or the app, or both. During the same period, the app identified 35 contacts that were also detected by conventional contact tracing and were assessed to be close contacts. Assuming the same percentage of app use among contacts as among cases of 22%, 236 of the total 1,073 close contacts could be expected to have been using the app and thus be potentially identified by the app during this period, resulting in an estimated sensitivity of 35/236=15%.
<sup>1</sup> As determined by conventional contact tracing done by public health staff.
<sup>2</sup> Excluding the 17 close contacts identified only by the app.
<sup>3</sup> Estimated for time period 1 August to 4 November 2020.

The 79 app-suggested contacts who met the close contact definition originated from 20 cases (Figure 1), and represented 0·3% (79/25,300) of all close contacts in NSW during the study period. Of these 79 close contacts, 62 (78%) were also identified by conventional contact tracing, leaving the additional yield of close contacts identified only via the app at 17, or 0·07% of all close contacts. These 17 contacts originated from four app-using cases (Figure 1), and all completed their quarantine period without becoming a case, hence no public exposure event to SARS-CoV-2 was prevented by the app during the study period. One or more of the following applied for all these 17 contacts: exposures had been forgotten or incorrectly recalled by cases; contact lists of venues visited by cases were unavailable or incomplete; and a broader close contact definition was sometimes applied by public health staff when multiple transmissions were known to have occurred in a specific setting.

The qualitative study component showed that the app was perceived as considerably less useful than public health staff had hoped. Its interface was not considered to be user-friendly, and especially in the beginning PHU staff required substantial assistance from central-level NSW Ministry of Health officials to access and interpret the data. This resulted in inefficiencies and delayed the roll-out of the app in NSW more generally. Public health staff reported that the process of ascertaining which of the app-suggested contacts that were not already through identified through the case interview were close contacts could be lengthy, which resulted in occasions of delay to notify close contacts, in particular when the number of app-suggested contacts was high. According to interviewees, the app did not shorten the timeframe for identifying close contacts. Some staff were also concerned that the app did not work equally reliably on all types of smartphones, with the number of contacts on iPhones being vastly underestimated while those on Android phones being overestimated. There was a general view that the app performed poorly when running in background mode. Overall, staff assessment of the app’s impact ranged from ‘not impacting much’ to being an additional step that increased workload without delivering any added value. The overwhelming response was that the app was not useful in contact tracing.

## DISCUSSION

COVID-19 is the first epidemic in which digital proximity tracing has been widely implemented. To our knowledge, the study reported here is the first large-scale assessment of the effectiveness and usefulness of a centralised digital tracing application. Our main findings over a 6-month period in Australia’s most populous state were that: less than a quarter of cases were using the app; app data from most app-using cases could not be used for contact tracing; the majority of app-suggested contacts were not close contacts; the majority of close contacts identified by conventional contact tracing were not detected by the app; and the additional close contacts identified by the app who would have been missed using only conventional contact tracing represented just 0·07% of the 25,300 close contacts recorded during the study period, none of whom presented an exposure risk to the general public.

We identify three broad issues that prevented the app from making a meaningful contribution to COVID-19 contact tracing in NSW. First, the uptake of the app among the population at risk of infection was low. While about 44% in a representative sample of the adult Australian population from June 2020 reported having downloaded the *COVIDSafe* app,^(19)^ in our study this figure was less than a quarter (22%) among those who actually contracted SARS-CoV-2 infection. Given that uptake levels above 50% are considered necessary for proximity tracing applications to help contain COVID-19,^(6)^ it is likely that its low uptake alone prevented the *COVIDSafe* app from making a substantial contribution to the COVID-19 response. Data security concerns, namely lack of privacy and fearing the normalization of governmental tracking; functionality issues, in particular negative impact on phone performance through increased battery usage; and privacy considerations due to the need for a centralized database, which is an integral design feature of tracing applications based on the centralized approach, were identified early as a potential barrier for acceptance of the *COVIDSafe* app.^(19-23)^

Second, with a positive predictive value below 40% and an estimated sensitivity of 15%, the diagnostic performance of the app was not sufficiently high to add value for COVID-19 contact tracing in NSW. Given that the app is intended to complement rather than to replace conventional contact tracing, a modest sensitivity might have been acceptable, particularly since some exposure settings were judged to be high risk and broader close contact criteria were applied. However, the app’s positive predictive value to correctly identify close contacts remained low despite filtering for Bluetooth signal exchange patterns that approximated the close contact definition. This meant that individual risk assessment by public health staff of all app-suggested contacts was necessary, as the imposition of unwarranted 14-day quarantine for the 60% of app-suggested contacts who were not deemed close contacts would have been unacceptable in a relatively low-transmission setting like NSW in 2020. Investigators of a small COVID-19 cluster that occurred in Sydney in March and April 2020 prior to the introduction of the *COVIDSafe* app speculated that the app might have helped to speed up the identification of contacts and links between cases.^(24)^ However, the poor diagnostic accuracy parameters established in our study suggest otherwise. The potential impact of the *COVIDSafe* app was also over-estimated in an earlier modelling study that projected a 25% reduction in cases if 27% of the population, i.e. slightly more than the proportion among cases in our study, had downloaded it.^(25)^ The only other empirical evaluation of a proximity tracing applications of a design similar to the *COVIDSafe* app was a small proof-of-concept study of Singapore’s *TraceTogether* app among staff and patients in a clinical setting. It found performance parameters even lower than in our study.^(26)^

Third, the app had a low perceived usefulness by public health staff while generating high workload. In particular, interviewed staff flagged the discrepancy between iPhones and Android phones in their ability to detect contacts. The failure to register contacts on iPhones was also noted in a pilot study of a similar app in the United Kingdom,^(27)^ which was later abandoned in part due to this problem.^(28)^ Another technical flaw of the app that affected its usefulness according to interviewed staff was its poor ability to register contacts unless the app was open. Problems with the *COVIDSafe* app picking up Bluetooth signals from locked phones or when running in the background had been noted early, but were not be sufficiently resolved.^(29)^ At the same time, the app did pick up a high proportion of contacts that were not deemed close contacts, which caused high workload for public health staff during the risk assessment process.

There were several limitations in our study. First, while app usage among cases in our study was only half of that estimated in the general population in Australia,^(19)^ we did not investigate the reasons for non-uptake among cases. Second, we could not determine to what extent the absence of registered contacts during the entire infectious period of some app-using cases was real or a result of technical problems of the app as described above. Third, prior to August 2020 the quality of surveillance data was insufficient to establish a reliable count of close contacts per case, and app use among contacts that were not app-suggested was not ascertained. As a result, we had to assume the same levels of app uptake among cases and contacts in order to estimate sensitivity (see footnote of Table 2). Fourth, we could not quantify the overall workload generated by the app for public health staff, in particular for the risk assessment of app-suggested contacts that did not meet the close contact definition.

Key implications of our findings are that conventional case interviews should remain the primary source of contact tracing information, and that all potential contacts, whether app-suggested or not, should undergo the same risk assessment by public health staff to establish their infection risk. The app may have shown greater benefits if uptake among the population at risk of SARS-CoV-2 infection had been higher. It may also have been more effective had it been used in a more targeted approach, for example for cases unable to give a reliable history, or for high risk exposure venues with incomplete attendance record keeping.

The costs for developing and operating the *COVIDSafe* app have been estimated at AUD 6·75 million until January 2021, with ongoing monthly maintenance costs of about AUD 100,000.^(30)^ These costs, coupled with the substantial additional workload required for public health staff to identify a very small number of additional close contacts, call for a formal cost benefit assessment of the *COVIDSafe* app, and of digital tools in the public health response to COVID-19 more generally.

In conclusion, the *COVIDSafe* app was not sufficiently effective to make a meaningful contribution to COVID-19 contact tracing in Australia’s most populous state during a 6-month period in 2020. Key issues were low uptake of the app, its poor positive predictive value and sensitivity, and difficulties for public health staff in accessing app-derived data. The additional contact yield was minimal and did not prevent any public SARS-CoV-2 exposure. At the same time, the app generated a substantial workload for public health staff, leading to high opportunity costs. Our findings contrast optimistic projections from modelling studies about the added value of digitally supported contact tracing. Further technical improvements coupled with comprehensive effectiveness evaluations are needed to yield real benefits of this technology for public health.

## Data Availability

Data produced in the present study are available upon reasonable request.

